# Knockdown AHNAK2 inhibits the progression of gastric cancer via the Wnt/β-catenin signaling pathway

**DOI:** 10.1101/2025.05.29.25328610

**Authors:** Xiang Liu, Lei Ma, Rui-Xiao Wang, Qi-Lun Liu

## Abstract

**Purpose:** AHNAK2 has been reported as tumor promoting protein by mediating tumor cell invasion and metastasis in a variety of malignancies, but the role of AHNAK2 in GC is indistinct.

**Methods:** Immunohistochemistry(IHC)was used to verify in GC tissues, clinical and pathological files were collected to figure out the correlation of AHNAK2 and prognosis in GC patients. GC cell lines were cultured to detect the expression and location of AHNAK2 by westernblot and immunofluorescence. Knockdown AHNAK2 to observe the invasion and metastasis of GC cell. Transcriptome sequencing was performed to explore the potential regulatory mechanism of AHNAK2.

**Results:** IHC results shows that AHNAK2 is upregulated in GC patients compared to normal people, overall survival in highly expressed AHNAK2 is poor,and AHNAK2 expression was positively correlated with lymph node metastasis. The expression of AHNAK2 in tumor cells is higher than in normal cell. Knockdown AHNAK2 decreased the proliferation, invasion, migration ability of GC cells, and increased cell apoptosis. Transcriptome sequencing reveals that AHNAK2 mediates GC progression by regulating Wnt/β-catenin axis.

**Conclusion:** Our findings suggest that AHNAK2 may promote the progression of GC by regulating Wnt/β-catenin signaling pathway.

## INSTRUCTION

Gastric Cancer (GC) has been a significant health concern globally, especially in East Asian countries^[1,2]^. In China, the incidence of GC morbidity is significantly high. According to statistics^[3]^, in 2020, the incidence and mortality of gastric cancer in China accounted for 43.9% and 48.6% of new cases and deaths in the world, respectively.

The GC or gastric adenocarcinoma is a highly complex heterogeneous malignant tumor^[4]^, which significantly limits the application of individualized treatment, thereby resulting in less than 30% of 5-year survival rate in advanced GC patients^[5]^. Although targeted therapeutic drugs such as herceptin have been clinically used^[6,7]^, surgery and perioperative chemotherapy remain the conventional therapy for advanced GC^[8]^. However, GC patients can acquire chemotherapy resistance, which reduces the therapy’s efficiency in inhibiting rapid growth and tumor metastasis ^[9]^. Therefore, studies on the potential molecular mechanisms of GC are urgently required to inhibit its progression.

AHNAK2 is a carcinogenic protein that has been observed to be associated with metastasis of tumors such as clear cell renal cell carcinoma (ccRCC)^[10]^, pancreatic ductal adenocarcinoma^[11]^, lung cancer^[12]^ and thyroid cancer(TC)^[13]^. Studies have shown that AHNAK2 is highly expressed in lung adenocarcinoma and is significantly associated with poor prognosis^[14]^. Knockout of AHNAK2 significantly reduces scratch healing ability. In addition, mechanism studies suggest that AHNAK2 may be involved in invasion and metastasis of lung adenocarcinoma through the AKT pathway, suggesting that AHNAK2 is involved in the pathogenesis and development of lung adenocarcinoma. However, the biological function of AHNAK2 and its potential regulatory mechanisms in GC remains undetermined.

Therefore, this study evaluated the expression and location of AHNAK2 by immunohistochemical analysis of 84 GC patients. Furthermore, the effect of AHNAK2 on prognosis was assessed by analyzing AHNAK2 expression and the patient’s clinicopathological features. Moreover, AHNAK2 expression in GC cell lines as well as the malignant biological behaviors of GC while knockdown AHNAK2, such as proliferation, migration, invasion, and apoptosis were assessed. Finally, transcriptome sequencing and further experiments verified the mechanism of AHNAK2’s possible involvement in the biological process of GC. This study provides a novel therapeutic approach and foundation for the clinical treatment of GC.

## MATERIALS AND METHODS

### Patients Samples and cell culture

This study included 84 radical gastrectomy-diagnosed primary GC patients who were admitted at the Cancer Hospital of Ningxia Medical University General Hospital from January 1, 2017, to December 31, 2017. The Ethics Committee of Ningxia Medical University General Hospital approved this research and the data were analyzed anonymously. All methods were performed in accordance with the relevant guidelines and regulations.

The human GC cell lines (AGS, HGC-27, MGC-803, and MKN-45) and corresponding normal gastric epithelial cells (GES-1) were purchased from the Chinese Academy of Medical Sciences (Shanghai, China) and cultured in RPMI-1640 medium augmented with FBS (Gibco, USA) and 1% penicillin-streptomycin (Gibco) incubating in 5% CO2 at 37°C. AHNAK2 expression was detected in all GC cell lines, and the one with the highest expression of AHNAK2 was selected for further experiments.

### Data profile collection

The patient’s clinicopathological and general data (gender, age, etc.) were retrospectively collected by His system. Furthermore, all the patients were followed up by telephone until December 31, 2022. The follow-up period was 3 - 60 months with a median of 47 months. There were 28 deaths, 25 survivors, and 5 lost to follow-up. The patient’s prognosis was expressed by the overall survival time.

### Immunohistochemistry

The cancer and adjacent tissues were submerged in paraffin, sliced (4 μm thick), baked, dewaxed, dehydrated, repaired using repair solution for 20 min, cooled down to room temperature, washed with phosphate-buffered saline (PBS), treated with endogenous peroxidase blocker, and then incubated in the dark for 10 min. The samples were then washed with PBS, probed with mono-antibody against AHNAK2 (1: 1000; Wuhan Sanying Company) at 4℃ overnight, rinsed with PBS, treated with horseradish peroxidase reductase working solution (Zhongshan Jinqiao Company) in the dark for 30 min, and rinsed again with PBS. The samples were then treated with 3, 3-diaminobenzidine (DAB) for color development, washed with distilled water, stained in hematoxylin for 5 min, and differentiated using hydrochloric acid alcohol. After washing, the samples were placed in tap water for anti-blue staining, dehydrated, and sealed using transparent neutral resin.

### Cell transfection

Sh-AHNAK2 and negative Control (sh-NC) were purchased from Hanheng Biotech Co., LTD (Shanghai, China). MKN45 cells were cultured in the 6-well plate until they reached 80% confluency. The media was then discarded, and Lipofectamine 3000 reagent was added for cell transfection. The media was refreshed after 24 h. After 48 h, cells with MOI = 30 were observed and selected for follow-up experiments. Transfection efficiency was detected by qRT-PCR and western blot analyses.

### Cell immunofluorescence staining

Cell lines with good growth status were cultured, digested, centrifuged, re-suspended, and cultured overnight in pre-treated small discs at appropriate concentrations. The cells were then rinsed with PBS, fixed, blocked, and probed with the primary antibody (1: 40 dilution) at 4℃ overnight. The next day, samples were rinsed with PBS buffer, treated with fluorescently labeled secondary antibodies (1:500 dilution) at room temperature for 2 h, rinsed again with PBS, and sealed with an anti-fluorescence quencher containing DAPI and blown dry. Based on the fluorescent attached to the secondary antibody, the expression of AHNAK2 was observed and photographed under the microscope through a suitable channel.

### Cell Proliferation Assay

Briefly, the transfected cells were cultured in a 6-well plate. When the number of cloned cells exceeded 50, the medium was aspirated, cells were rinsed with PBS thrice, fixed with 4% paraformaldehyde for 15 min at room temperature, washed again with PBS, and stained with 0.5 mL 0.05% crystal violet solution for 20 min. The excess crystal violet dye was cleaned by rinsing with PBS thrice, and cells were air-dried before clone formation rate analysis and imaging via a digital camera.

### Cell Migration and Invasion Assay

Cell migration and invasion abilities were evaluated by wound healing and transwell analyses, respectively. For wound healing or migration analysis: the cells were cultured in a 6-well plate at 37°C for 8 h. Upon 80% confluency per well, the cells were then scraped using a pipette tip to create a wound, washed with PBS, and allowed to grow in serum-free media. The wound closure distance was measured under a microscope at 0 and 48 h. For invasion or Transwell analysis: First, the matrix gel was melted and diluted in a serum-free medium at a ratio of 1:6, which was then added to the upper portion of the perforated chamber. In the lower chamber, 500 mL of serum-free medium was added to rehydrate the basement membrane, whereas in the upper chamber, 1 mL of cell suspension was then added for 24 h. The uninfected cells were discarded, and the remaining cells were washed thrice, fixed with 4% paraformaldehyde, and stained with 0.1% crystal violet. The detection of migration ability was explored in the absence of matrix gels.

### Flow Cytometry Assay

The cell apoptosis rate was determined by following the instructions provided in the Cell Apoptosis Detection kit. Briefly, the transfected MKN-45 cells were re-suspended and then treated with 10 mL Annexin V-fluorescein isothiocyanate (FITC) and 5 mL propidium iodide (PI) for 15 min at 25°C in the dark. Subsequently, the apoptotic rate was analyzed using a flow cytometer (BD Biosciences, San Jose, CA, USA).

### qRT-PCR

The tumor cell RNA was extracted per the instructions of the kit (Invitrogen; Thermo Fisher Scientific, Inc) and then reverse transcribed into cDNA, which was then subjected to qRT-PCR. The amplification conditions were as follows: 95°C for 5 min, followed by 40 cycles of 95°C for 30 s, 55°C for 20 s, and 72°C for 20 s. The relative expression of AHNAK2 was analyzed using the 2-ΔΔCT method. GAPDH was used as a housekeeping gene. The sequences of primers employed for qRT-PCR are as follows:

AHNAK2:

Forward: 5’-GACCTGCCTCTTCCCAAACA-3, Reverse: 5’-GCGAGTACTTGGTCATGGCT-3.

GAPDH:

Forward: 5’-TCAAGGCTGAGAACGGGAAG-3, Reverse: 5’-TCGCCCCACTTGATTTTGGA-3.

### Western Blotting

The cultured cells were treated with 100 μL protein lysate, scraped, and transferred to the EP tube on ice for 30 min. The cells were then centrifuged at 4℃ and at 12000 rpm for 15-30 min to collect the supernatant. The acquired proteins in the supernatant were quantified using the BCA method. The proteins (30 μg) were then subjected to gel electrophoresis, transferred to the membrane, blocked with 5% skim milk for 2 h, rinsed with TBST buffer thrice for 5 min each time, and then treated overnight with polyclonal antibody against rabbit and human AHNAK2 (1: 1000; American OriGene Company) and β-actin (Wuhan Sanying Company) at 4℃. The next day, the membranes were rinsed with TBST buffer, probed with the secondary anti-sheep and anti-rabbit HRP antibodies (Wuhan Sanying Company) for 2 h at room temperature, washed with TBST buffer, and reacted with ECL for protein band development. Image J was used to analyze the gray value of the image and the relative expression level of the AHNAK2 protein. The experiment was repeated three times.

### Statistical Analysis

All data were analyzed by SPSS 22.0 and GraphPad Prism 8 and presented as mean ±standard deviation (SD). The intergroup differences were compared by T-test, whereas the difference between more than 2 groups was compared by one-way analysis of variance. Kaplan-Meier curves were used to estimate the patient’s survival rate. *p* < 0.05 indicated that the data had statistically significant differences.

## RESULT

### AHNAK2 is upregulated in GC tissues

This study performed immunohistochemical staining of samples from 84 GC patients to assess the expression and localization of AHNAK2. The results indicated that 69.0% (58/84) GC patients had high AHNAK2 expression in the tumor tissue, while 31.0% (26/84) GC patients had no AHNAK2 expression, which was also negative in all adjacent normal tissues. Altogether, AHNAK2 was observed to be upregulated in GC tissues ( Figure 1A).

**Figure 1:**
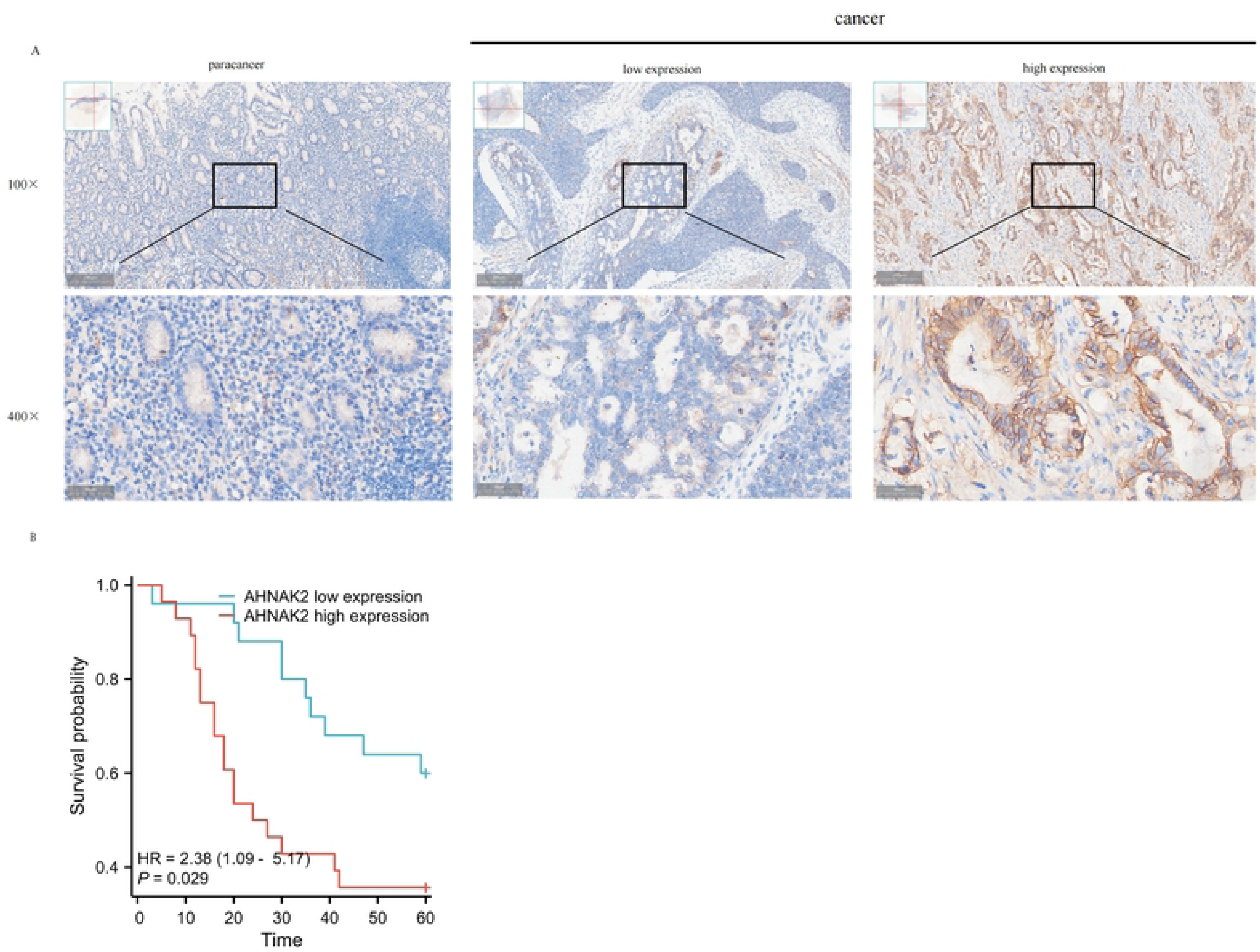
AHNAK2 is upregulated in GC and associated with poor prognosis. (A) Expression of AHNAK2 in gastric cancer and adjacent tissues. (B) AHNAK2 is associated with poor prognosis of gastric cancer.

### Relationship between AHNAK2 and clinicopathological features of GC patients

The 58 AHNAK2-positive patients were categorized into high and low AHNAK2 groups based on expression levels acquired by multiplying the staining degree score (percentage of positive cells) with the staining intensity score, with a median of 2 as the limit. There were 32 cases with high AHNAK2 expression and 26 cases with low expression. It was also observed that AHNAK2 expression was positively correlated with lymph node metastasis (LNM) in GC patients (*p*<0.05). Furthermore, AHNAK2 was not associated with age, sex, tumor differentiation, pT stage, pathological stage, and the presence of vascular cancer thrombus (*p*>0.05) (Table 1).

**Table 1:** Correlation of AHNAK2 expression and clinicopathological features in GC patients.

| Characteristic | Total | AHNAK2 expression |  | *P* value |
| --- | --- | --- | --- | --- |
|  |  | Low | high |  |
| Gender |  | 26 | 32 | 0.431 |
| male | 44 | 21 | 23 |  |
| female | 14 | 5 | 9 |  |
| Age(years) |  |  |  | 0.771 |
| <60 | 30 | 14 | 16 |  |
| ≥60 | 28 | 12 | 16 |  |
| Differentiation |  |  |  | 0.329 |
| Well | 8 | 4 | 4 |  |
| Moderate | 24 | 10 | 14 |  |
| poor | 26 | 12 | 14 |  |
| pT stage |  |  |  | 0.051 |
| T1 | 3 | 3 | 0 |  |
| T2 | 10 | 7 | 3 |  |
| T3 | 11 | 4 | 7 |  |
| T4 | 34 | 12 | 22 |  |
| pN stage |  |  |  | 0.032 |
| N0 | 10 | 8 | 2 |  |
| N1+N2+N3 | 48 | 18 | 30 |  |
| TNM Stage |  |  |  | 0.258 |
| I - II | 20 | 11 | 9 |  |
| III-IV | 38 | 15 | 23 |  |
| Vascular invasion |  |  |  | 0.272 |
| Absent | 33 | 19 | 14 |  |
| Present | 17 | 7 | 10 |  |

The 58 AHNAK2-positive GC patients were followed up till December 31, 2022 (5 years postoperatively). During this period, 5 patients were lost to follow-up and 28 died, with an overall survival rate of 47.2%. Among these, the 1-, 3-, and 5-year survival rates of high AHNAK2 expression GC patients were 82.1% (23/28), 42.9% (12/28), and 35.7% (10/28), respectively. Whereas the 1-, 3-, and 5-year survival rates of low AHNAK2 expression GC patients were 95.7%(22/23), 69.6%(16/23), and 56.5%(13/23),respectively. Overall, these data indicate that high AHNAK2 expression patients had a lower survival rate than low AHNAK2 expression patients (*p*<0.05) (Figure 1B).

### AHNAK2 was up-regulated in GC cell lines

The normal gastric epithelial cells (GES-1) and four GC cell lines (HGC-27, AGS, MGC-803, MKN-45) were selected to analyze their AHNAK2 expression. It was revealed that AHNAK2 expression in all GC cells was higher than in GES-1 cells, with MKN-45 GC cells indicating the highest expression (Figure 2A-D). Therefore, MKN-45 GC cells were selected for subsequent experiments. Furthermore, AHNAK2 was observed to be predominantly expressed on the cell membrane. From these results, it was inferred that AHNAK2 was upregulated in GC cells.

**Figure 2:**
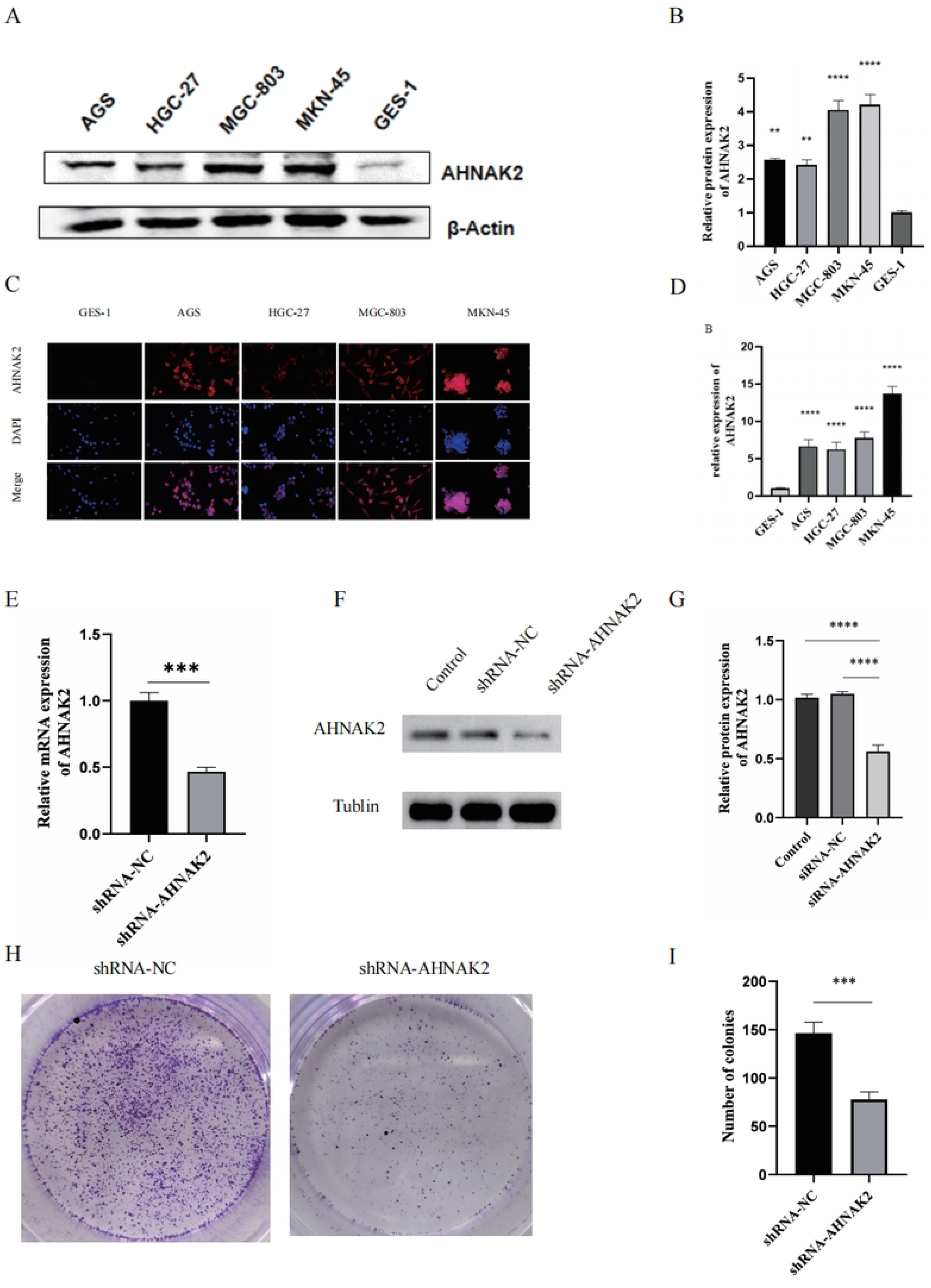
AHNAK2 promotes the proliferation of GC cells. (A) Western blot showed the expression of AHNAK2 in different GC cell lines compared with normal cell. (B) Semi-quantitative analysis of AHNAK2 expression in different cells in Fig2A. (C) immunocytofluorescent analysis of AHNAK2 in different cells. Scale=100 μm. (D) Semi-quantitative analysis of AHNAK2 expression in different cells in Fig2C. (E) Knockdown efficiency of shRNA-AHNAK2 by qRT-PCR.(F) Western blot was used to detect the expression of AHNAK2 in different groups. (G) Semi-quantitative analysis of AHNAK2 expression in different cells in Fig2F. (H) Colony formation assay of GC cells in shRNA-AHNAK2 and shRNA-NC groups.(I) Semi-quantitative analysis of Fig2G. \*\**p*< 0.05; \*\*\**p*< 0.001, \*\*\*\**p*< 0.0001.

### AHNAK2 knockdown inhibits MKN-45 cell proliferation

To further study the biological role of the AHNAK2 in GC, MKN-45 cells were utilized to construct stable AHNAK2 knockdown cell lines via lentivirus-mediated gene transfection. qRT-PCR analysis revealed that the knockdown efficiency of mRNA level in shRNA-AHNAK2 cells was 53% (Figure 2E). Furthermore, western blot analysis also confirmed the construction of AHNAK2 knockdown stable transmutation (Figure 2F-G). Cell clonogenesis experiments are used to assess cell proliferation capacity and population dependence to indicate differences in cell-independent viability. The cancer cell’s malignant degree increases with the increase in clonogenesis ability. Here, the clonogenesis assay was performed to assess how AHNAK2 downregulation affects the malignant proliferation of GC cells. It was revealed that on the 13th day after shRNA-AHNAK2 transfection, the number of clone cells was significantly reduced compared with that in the shRNA-NC group, indicating that AHNAK2 knockdown could inhibit GC cell proliferation (Figure 2H-I).

### AHNAK2 knockdown suppresses MKN-45 cell migration and invasion, as well as induces apoptosis

The cell’s ability to migrate is the premise of tumor invasion and metastasis. To assess the cell’s migration ability, the shRNA-NC and shRNA-AHNAK cells were scratched, cultured, and photographed at 0 and 48 h, and the relative migration area of the cells transferred to the scratch zone under each high-power field of view was calculated after 48 h. Compared with shRNA-NC, the migration ability of the shRNA-AHNAK2 cells was significantly decreased (*p* < 0.05) (Figure 3A-B). The transwell assay was performed to assess the effect of AHNAK2 knockdown on the GC cell’s invasion ability. Compared with the control and shRNA-NC cells, the invasion ability of the shRNA-AHNAK2 cells was significantly inhibited. This suggests that AHNAK2 can promote the invasion of GC cells (Figure 3C-D). Furthermore, the flow cytometry analysis revealed that the apoptotic rate of the shRNA-AHNAK2 cells was significantly higher than the MKN-45 and shRNA-NC cells (Figure 3E-F, *p*<0.05), indicating that AHNAK2 knockdown could promote GC cells apoptosis. Moreover, western blot analysis was performed to assess the levels of anti-apoptotic protein Bcl-2 and pro-apoptotic proteins Bax and Caspase3. It was found that the expression levels of apoptotic proteins were increased after AHNAK2 knockdown (Figure 3G-H).

**Figure 3.**
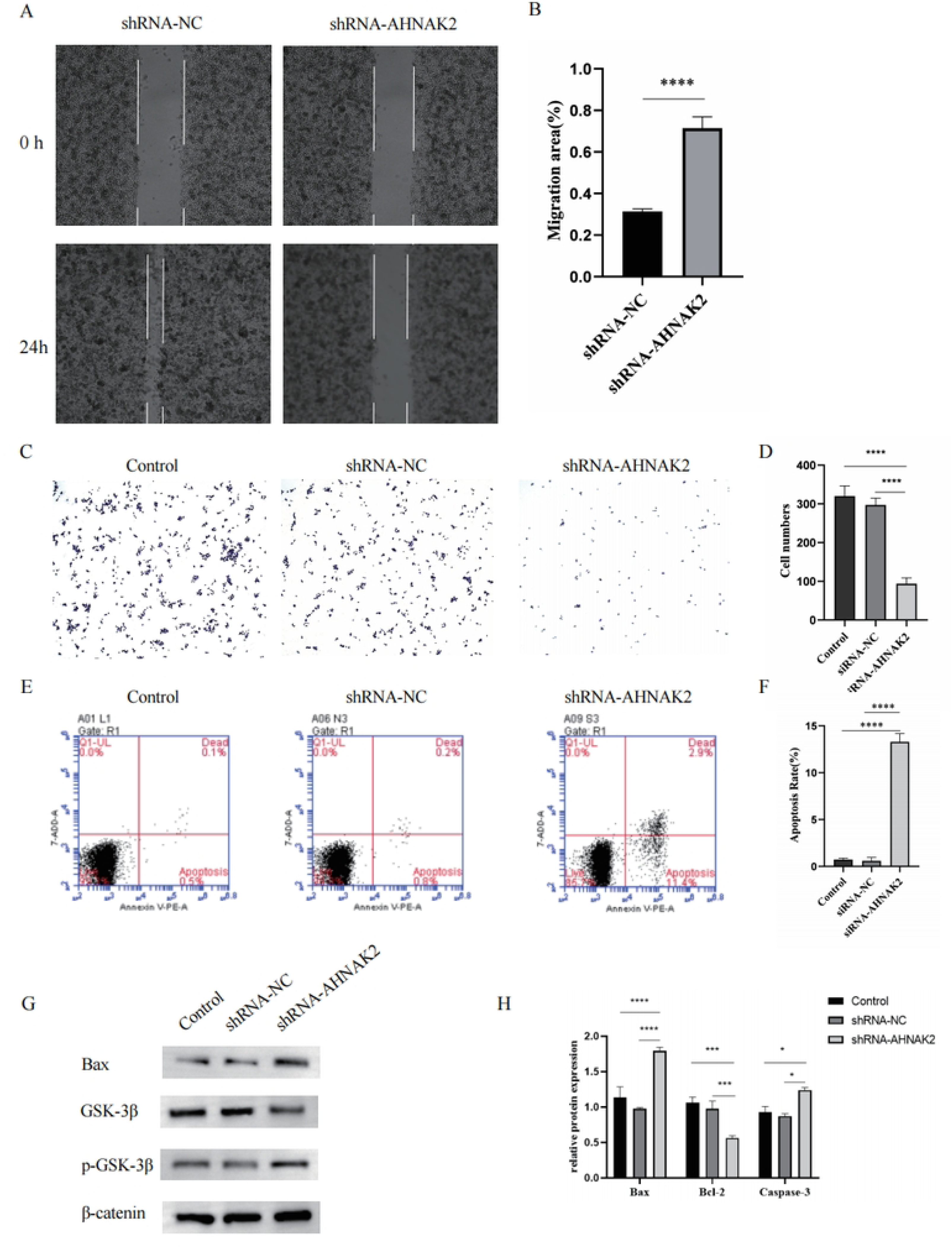
Effects of AHNAK2 on invasion, migration, and apoptosis of GC cells.(A) Cell migration assay was performed at 0 hour and 48 hour in different groups.(B) Semi-quantitative analysis of Fig3A. (C) Transwell chamber invasion assay was used to detect the invasion ability of cells in different groups. (D) Semi-quantitative analysis of Fig3C.(E) Flow cytometry was used to detect cell apoptosis in different groups.(F) Semi-quantitative analysis of Fig3E.(G) western blot was used to detect the expression of Bcl-2、 Bax and Caspase3 protein expression in MKN-45 cells after knocking down AHNAK2.(H) Semi-quantitative analysis of Fig3G.\**p* < 0.05; \*\*\**p* < 0.001, \*\*\*\**p* < 0.0001.

### AHNAK2 knockdown inhibits the progression of GC cells by suppressing the Wnt/β-catenin signaling pathway

To explore the potential molecular mechanism of AHNAK2 in regulating GC progression, transcriptional sequencing, and bioinformatics analysis were performed on the shRNA-NC and shRNA-AHNAK2 cells, which revealed 790 up-regulated and 1389 down-regulated differentially expressed genes (DEGs) between the two groups (Figure 4A). Furthermore, KEGG and GSEA enrichment analysis of these DEGs showed that AHNAK2 might be associated with GC progression via the Wnt/β-catenin signaling pathway (Figure 4B - C). To investigate whether AHNAK2 regulates GC progression by mediating Wnt/β-catenin, the expression of key Wnt/β-catenin pathway proteins including Wnt3a, GSK-3β, p-GSK-3β, β-catenin, PCNA, cyclinD2, Cdk4, and c-Myc in control, shRNA-NC and shRNA-AHNAK2 cells was assessed by Western blot. It was found that AHNAK2 knockdown decreased the expressions of Wnt3a, GSK-3β, p-GSK-3β, β-catenin, PCNA, cyclinD2, Cdk4, and c-Myc (Figure 4D-F, p < 0.05). Western blot verification results were consistent with the above transcriptome sequencing results, indicating that AHNAK2 may play an effect on GC via the Wnt/β-catenin pathway.

**Figure 4.**
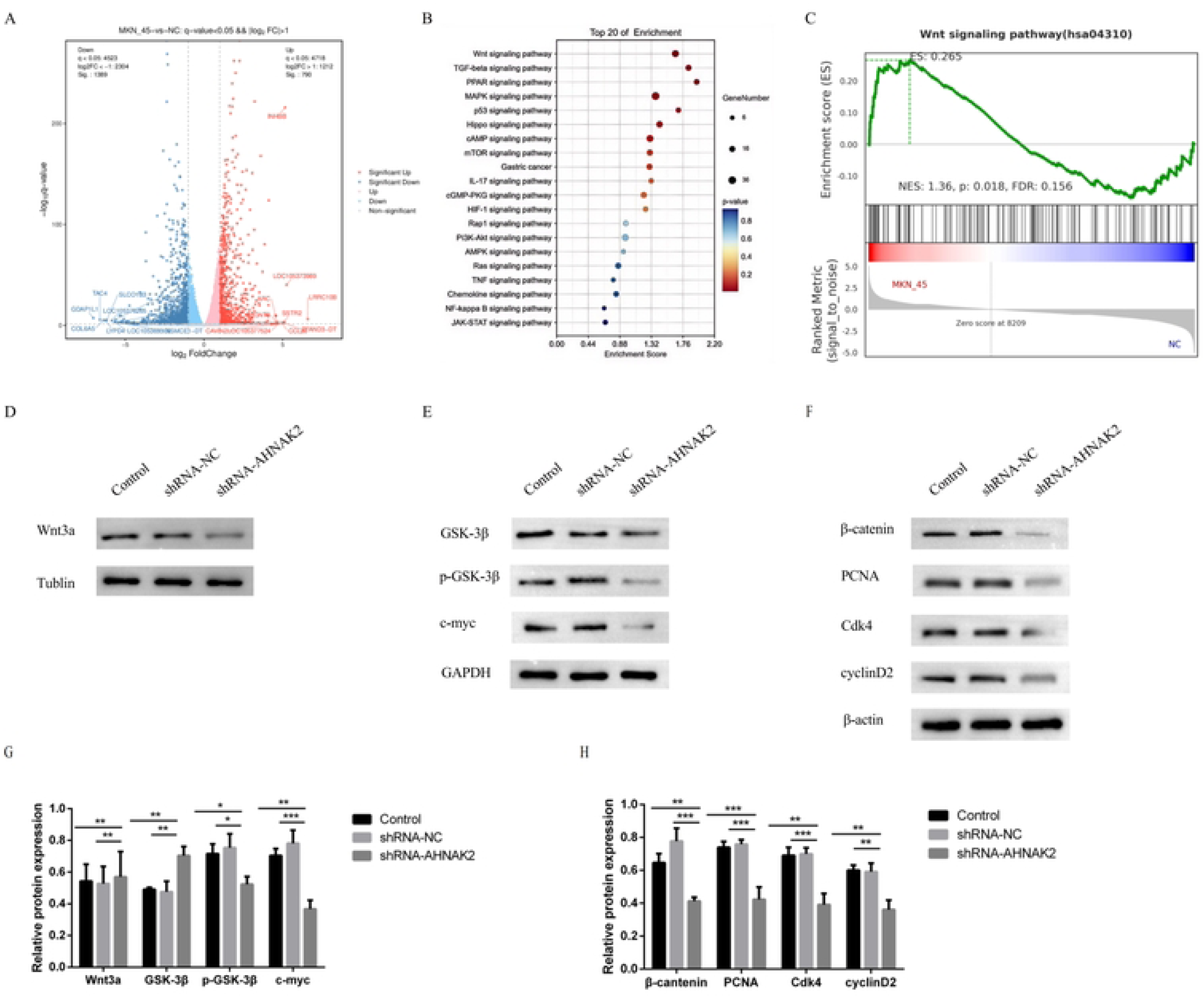
AHNAK2 promotes the proliferation, migration and invasion of gastric cancer by mediating Wnt/β-catenin signaling pathway.(A) Differential genes in cell sequencing data from shRNA-NC and shRNA-AHNAK2 groups of GC cell are shown in volcano plots. Red dots indicate up-regulated genes, blue dots indicate down-regulated genes. (B) KEGG enrichment showed the top 20 signaling pathways of differentially expressed genes. (C) GSEA analysis showed that Wnt/β-catenin signaling pathway was enriched between the two groups. (D) Western blot was used to detect the expression of Wnt3a in different groups.(E) Western blot was used to detect the expression of GSK-3β, p-GSK-3β and c-myc in different groups.(F) Western blot was used to detect the expression of β-catenin,PCNA,Cdk4 and cyclinD2 in different groups.(G)Semi-quantitative analysis of Fig4D-E. (H) Semi-quantitative analysis of Fig4F. \**p* < 0.05, \*\**p* < 0.01; \*\*\**p* < 0.001.

## DISCUSSION

Gastric cancer is an epithelial malignancy with the 4th highest mortality rate among all cancers globally^[3]^. Due to its latent early symptoms, most patients are diagnosed at an advanced stage. It has been estimated that the 5-year survival rate of GC patients with distant metastasis is less than 5%^[15]^. Currently, the GC treatment mainly includes surgery, radiotherapy, and chemotherapy, which effectively improve the patient’s survival rate and quality of life, however, there are still cases of recurrence, metastasis, and drug resistance^[16]^. Although several studies have been performed on the etiology and pathophysiology of GC in recent years, the reliable and efficient therapeutic targets are limited.

AHNAK2 is a member of the AHNAK family that is located on human chromosome 14q32^[17]^.It has been indicated that AHNAK2 is crucially associated with the progression of various cancers^[18]^. A study analyzed 178 pancreatic ductal adenocarcinoma patients and indicated that high expression of AHNAK2 was associated with poor prognosis^[19]^. High AHNAK2 expression was also found to be upregulated in lung adenocarcinoma, and its high expression was positively correlated with LNM, severe staging, and poor survival^[20,21]^. Another study indicated that AHNAK2 plays a carcinogenic role in ccRCC, and high AHNAK2 expression has been linked with late-stage, metastasis, and shorter survival rate^[10]^. Furthermore, Koguchi^[22]^ revealed that AHNAK2 expression was high in bladder cancer, whereas, the normal uroepithelial cells no had no AHNAK2 expression. Moreover, their survival analysis indicated that high AHNAK2 expression was linked with poor prognosis. In this study, high AHNAK2 expression was observed in both GC tissues and cell lines, and it was positively correlated with LNM in GC patients. Furthermore, high AHNAK2 expression patients had poor prognosis, suggesting that AHNAK2 may be involved in tumor progression and therefore, AHNAK2 can be utilized as an indicator for poor prognosis in GC patients.

Metastasis of cancer cells is a complex biological process, which involving invasion, migration and proliferation^[23]^. Those processes not only greatly affect outcomes and survival expectations of patients, but also make treatment strategies tricky. To date, the biological role of AHNAK2 in cancer is relatively consistent. Studies have shown that AHNAK2 expression is up-regulated in TC tissues, especially in metastatic TC tissues, and down-regulation of AHNAK2 can inhibit the migration, invasion and metastasis of TC^[24]^. Lin also found that high AHNAK2 expression was closely related to the pathological staging of TC, whereas its knockdown inhibited the proliferation, metastasis, and epithelial-mesenchymal transformation (EMT) of thyroid cancer cells^[25]^. EMT is considered to be the premise of cancer growth, invasion and metastasis, and plays an important role in cancer progression and metastasis, EMT can also increase the resistance of cancer cells to chemotherapy and immunotherapy^[26–28]^. At the same time, studies have shown that AHNAK2 plays as tumor promoter by promoting epithelial cells which acquire a series of mesenchymal features^[14]^. Wang suggested that when AHNAK2 was down-regulated, MAPK/ERK signaling pathway was inhibited, and lung cancer cell migration and invasion abilities were reduced, whereas the apoptosis rate was significantly enhanced^[12]^. A study on the regulation of AHNAK2 in uveal melanoma found that reducing the expression of AHNAK2 could inhibit the proliferation, migration and invasion of uveal melanoma cells, and also inhibit the activation of the phosphoinositide 3-kinase(PI3K) signaling pathway^[29]^.Similarly, the current study suggests that AHNAK2 plays a tumorigenic role in tumor growth and progression in ccRCC. Knocking down AHNAK2 inhibits tumor proliferation, migration and colony formation in vitro and reduces tumor growth in vivo. In addition, downregulation of AHNAK2 inhibits fatty acid and lipid synthesis, which is an important process for maintaining cancer energy and cellular nutrition, suggesting that the carcinogenic effects of AHNAK2 may be achieved by altering cellular tumor metabolism^[10]^. Recent studies have found that AHNAK2 is also highly expressed in various adenocarcinoma tissues, while it is almost not expressed in normal glandular tissues. In vitro experiments found that knocking down AHNAK2 can significantly inhibit the proliferation and migration ability of adenocarcinoma cell lines, proving that AHNAK2 is a biomarker and it can be a potential therapeutic target for adenocarcinoma^[30]^. Here, our results demostrated that knockdown AHNAK2 can alleviate clonogenesis ability, which means malignant growth of GC cells was reduced. And at the same time, suppressing AHNAK2 can reduce the ability of migration and invasion of GC cells. while the apoptosis rate increased. Altogether, these results indicated that inhibiting AHNAK2 might suppressed the progresson of GC. This further confirms that AHNAK2 is involved in GC progression and plays an important biological role.

In the early stage of this study, it was found that AHNAK2 can promote GC progression. In order to further explore its potential regulatory mechanism, transcriptomic sequencing was performed. The enrichment analysis results suggested that the significant difference pathway was Wnt signaling pathway. Wnt signaling pathway is one of the main pathways involved in human embryo and organ development, it has been related to various biological processes of tumor cells, such as proliferation, migration, differentiation, and apoptosis^[31]^.The activation of Wnt signaling pathway is mainly divided into classical Wnt/β-catenin signaling pathway, Wnt/Ca2+ pathway and Wnt/PCP pathway^[32]^.Once the Wnt/β-catenin signaling pathway activated, the core protein molecule β-catenin is transferred from the cytoplasm to the nucleus, thus promoting the expression of related target genes such as cell proliferation, differentiation and migration^[33]^.More and more studies have confirmed that the Wnt signaling pathway plays an irreplaceable role in the development and progression of GC^[34–36]^. It has been estimated that 46% of gastric tumors have dysregulated Wnt/β-catenin pathway^[37]^. A study analyzed 13 GC cell lines and revealed nuclear localization of endogenous β-catenin and increased TCF/LEF transcriptional activity, confirming abnormal Wnt signaling^[38]^.The KEGG pathway analysis has revealed that the Wnt pathway is the 3rd active pathway in GC, and its downstream factors RNF43, AXIN1/2, CTNNB1, and APC are frequently mutated^[39]^. In our study, we found that the expression of related proteins of Wnt pathway was decreased while knockdown AHNAK2, suggesting that AHNAK2 may regulate GC progression via the Wnt/β-catenin pathway.

In summary, this study indicates that AHNAK2 participates in the regulation of malignant biological behaviors such as proliferation, apoptosis, migration and invasion of gastric cancer cells, and it may achieved by the activation of Wnt/β-catenin signal, revealing that AHNAK2 can be used as a potential therapeutic target for the treatment of gastric cancer. It provides a new theoretical mechanism and support for the treatment of gastric cancer.

## CONCLUSION

In summary, this study revealed that the AHNAK2 protein can serve as a biomarker to indicate poor prognosis in GC patients and a novel molecular target for diagnosis and treatment of this malignancy.

## Data Availability

Datasets generated and/or analyzed during the current study are available at the National Center for Biotechnology Information in the repository, [https://dataview.ncbi.nlm.nih.gov/object/PRJNA1214896?reviewer=o3idag4aqm507rh0obobeaei4r] ". All data and materials are available in the paper and the supplementary information. Source data are provided with this paper.

## Acknowledgments

The RNA libraries were sequenced by OE Biotech, Inc, Shanghai, China. We are grateful to OE Biotech, Inc, (Shanghai, China) for assisting in sequencing and/or bioinformatics analysis. All colleagues involved in this study for their contributions.

## Funding

This work is supported by grants from the National Natural Science Foundation of China (82060663), and the Natural Science Foundation of Ningxia (2020BEG03031).

## Data availability

Datasets generated and/or analyzed during the current study are available at the National Center for Biotechnology Information in the repository, [https://dataview.ncbi.nlm.nih.gov/object/PRJNA1214896?reviewer=o3idag4aqm507 rh0obobeaei4r] “. All data and materials are available in the paper and the supplementary information. Source data are provided with this paper.

## Authors’ contributions

Q-L L and XL designed the study and confirmed the authenticity of all the raw data. XL and LM performed data analysis and drafted the manuscript. XL and LM performed verification experiments and improved the language of the manuscript. R-X W contributed to technical support and histological evaluation. All authors have read and approved the final manuscript.

## Ethics approval and consent to participate

The study on human tissues was approved by the Ethics Committee of Ningxia Medical University General Hospital (ethics code: KYLL-2024-0023).

## Patient consent for publication

Not applicable

## Competing interests

The authors declare no competing interests.

## Notes

### Competing Interest Statement

The authors have declared no competing interest.

### Funding Statement

The author(s) received no specific funding for this work.

